# Parsing the Functional Heterogeneity of the Psychosis Spectrum: A Clinically Informed Application of Genomic Structural Equation Modeling

**DOI:** 10.64898/2026.09.08.26362224

**Authors:** Luca Sclisizzo, David AA Baranger, Margaret L Clapp Sullivan, Alexander S Hatoum, Marco Guidice Del, Emma C Johnson

## Abstract

Schizophrenia, bipolar disorder, and major depression vary in their age of onset, cognitive impacts, degree of impairment, and clinical course. However, since psychosis is a transdiagnostic feature that occurs across all three of these disorders, they can be represented as a spectrum. We leveraged genomic data to explore different functionally distinct components of the psychosis spectrum. We used Genomic Structural Equation Modelling (Genomic SEM) to define four factors indexing specific aspects of psychosis-related symptoms shared across depression, schizophrenia, and bipolar disorder: a general Psychosis factor with low Cognitive functionality (gPsychosisCognition), a specific Psychosis factor independent of cognition (sPsychosis), a specific Affective factor independent of psychosis (sAffective) and a specific Mania factor independent of depression (sMania). We then used Stratified Genomic SEM and transcriptomic analyses to characterize biological functions and temporal trends associated with these factors. sPsychosis, sAffective, and sMania exhibited differential genetic correlations with personality, with sPsychosis and sAffective showing positive correlations with Openness, while Neuroticism showed a strong positive correlation with sAffective and a negative correlation for sMania. GABAergic, cortical, and CA1-CA3 enrichments were significant across the continuum with evidence for factor specific enrichment: dentate gyrus, CA4 and cerebellum were non-significant only for sAffective and oligodendrocytes enriched only in gPsychosisCognition. Transcriptomic analyses revealed different temporal expression trends across factors, identifying a mostly prenatal expression for sMania and sAffective, and more consistent expression across development for sPsychosis. We demonstrated that a clinically informed genomic model can shed light on the functional heterogeneity of psychosis, identify both unique and shared biological components and highlight its partial neurodevelopmental origin.

## Introduction

Current diagnostic categories in psychopathology that are treated as separate disorders often show high degrees of comorbidity and shared genetic factors (1,2), prompting many researchers to build dimensional transdiagnostic models that describe broad spectra of correlated symptoms (3–5). At the same time, diagnostic categories are characterized by a striking degree of clinical heterogeneity that could arise from functionally different etiological processes (6,7) leading to variability in burden, onset, remission and relapse.

Psychosis is a feature shared across multiple diagnostic categories that exhibit substantial comorbidity and overlap. This similarity motivated scholars to group schizophrenia (SCZ), bipolar disorder (BD), the mixed condition of schizoaffective disorder (SZA) and other related conditions (such as delusional disorder and schizotypal personality disorder) into a single psychosis spectrum (8–11), or even a “superspectrum” that blends with the detachment symptoms of schizoid and avoidant personality disorders (12).

Despite their overlap, psychosis-spectrum disorders are considerably heterogenous in their severity, age of onset, and prognosis (Del Giudice 2018). However, different authors (9,10,13,14) have posited that this heterogeneity may be largely explained by a single gradient-like dimension of functionality that is (partially) independent of the severity of the clinical manifestation (15) (Figure 2A). The more dysfunctional pole of the gradient is marked by negative symptoms such as blunted affect, asociality and avolition, while the opposite and more functional pole is characterized by manic affective symptoms (13) (Figure 2A). This gradient-like dimension goes beyond the standard distinction between negative and positive symptoms, and accounts for the heterogeneous cognitive patterns observed in this clinical domain (9) along with age of onset, clinical course and genetic burden (10,13).

### Phenotypic Overlap and Heterogeneity in Psychosis

From the phenotypic standpoint, SCZ and BD show similarities in both symptoms and environmental risk factors. Perinatal complications, childhood difficulties and harsh rearing contexts have been linked as risk factors to both conditions (16). Cognitive impairments and lower-than-average IQ have been detected in both SCZ and BD, with SCZ showing greater compromission than BD (9,17). While cognitive impairment does not neatly discriminate between diagnoses (since a sizable overlap in IQ has been observed between SCZ and BD cases (18–20), SCZ is systematically associated with more severe brain atrophy (21,22), and lower rates of remission and recovery than BD. SCZ patients in remission show intermediate cognitive and functioning profiles compared to their acute counterparts and BD patients (23,24).

Within SCZ, cognitive functioning and clinical course are worse in early-onset cases with predominantly negative symptoms, and comparatively better in cases marked by predominant positive and/or affective symptoms (17,20,25–27). Vice versa, BD patients with delusions or hallucinations show lower IQ, earlier onset, and worse prognosis than those with only mood-related symptoms (9,28). Moreover, early-onset BD has a cognitive and neurodevelopmental profile similar to that of SCZ, supposing similar developmental factors across these conditions (29). The profile of SZA is intermediate between those of SCZ and BD, reflecting a mixture of psychotic and affective symptoms (9).

Psychotic symptoms are not limited to SCZ and BD, since cases of unipolar depression (MDD) can have psychotic features such as delusions and hallucinations (30); melancholic depression often co-occurs with BD and can be treated with antipsychotics (31–33). Moreover, psychotic MDD is more frequent in patients with family history of SCZ and BD and is more likely to evolve in BD SCZ than non-psychotic MDD (34,35). However, due to the high degree of heterogeneity of MDD and to the diagnostic difficulties of tracking psychosis in MDD (33,36), the role of unipolar depression (MDD) with respect to the psychosis spectrum remains less clear.

In short, the clinical heterogeneity that characterize the psychosis domain is not accounted by clear-cut distinctions between diagnostic categories, but rather by a sub-diagnostic continuous dimension explicitly posited by Craddock & Owen (13) and later reaffirmed by Lynham et al., (9).

### Genetic Overlap and Heterogeneity in Psychosis

The genetic literature provides crucial information about the gradient-like dimension of psychosis, and how it relates to the genetic architectures of the disorders. Firstly, studies consistently find substantial genetic correlations among SCZ, BD and MDD (2,11,37–39). As a result, there is a growing tendency to group them into hierarchies of broad and specific factors (2,11,40). Secondly, the phenotypic cognitive patterns observed across diagnoses are also maintained at the genetic level: SCZ shows larger and more consistent negative genetic correlations than BD with cognitive traits such as IQ, executive functions, and educational attainment (41–43). A parallel pattern is found within the diagnostic category of SCZ, whose subtypes show differential genetic correlations with cognitive and affective traits (44,45). Moreover, polygenic scores for improved cognitive ability have been found to predict better trajectories of clinical progression in patients with first-onset psychosis (46). Importantly, polygenic scores for BD and SCZ can cross-predict clinical features in the psychosis domain: BD polygenic score are predictors of manic symptoms in SCZ patients; on the other side, SCZ polygenic scores are associated with earlier onset of BD, increased psychosis features among BD cases and greater levels of negative symptoms in SCZ patients (47,48).

Regarding MDD, families and twin studies have found familiar co-occurrence of psychotic MDD with both SCZ and BD, with indications that psychotic MDD reflects co-occurring liabilities to SCZ and BD (34,49). Moreover, psychotic MDD shows a higher correlation with SCZ than its non-psychotic counterpart; MDD polygenic scores are associated with lower odds of psychotic MDD whereas SCZ, BD and BD-I polygenic scores predict increased odds of psychotic MDD (35). Thus, it is likely that the well-recognized heterogeneity of depression includes variants that may be better characterized as belonging to (or at least intersecting with) the spectrum of psychosis; vice versa, certain conditions within the spectrum (notably BD-II) show stronger clinical and genetic links with MDD than others (e.g., BD-I) (40,47,50).

In summary, the phenotypic pattern that motivated the gradient-like dimension perspective can be observed also at the genetic level, where the clinical and cognitive profiles of psychosis conditions are also reflected by genetic studies.

### Genomic Dissection of the Psychosis Spectrum

Previous studies have used different approaches to dissect the psychosis spectrum with the common aim to find homogeneous genetic sub-components (40,45,51,52). In one example, Mallard et al. (40) used an Exploratory Factor Analysis (EFA) with *promax* rotation on the genetic variance-covariance matrix to model two correlated factors, one with loadings from SCZ, SZA and BD-I and another with loadings from MDD, BD-II, and a variety of manic, depressive and psychotic symptoms. As expected, rotated EFA applied to genomic data resulted in two positively correlated factors (Mallard et al. 2022), broadly representing more common forms of psychiatric illness (MDD, BD-II) and rarer, more severe conditions (SCZ, BD-I).

In the current paper, we take a somewhat different perspective, deliberately focusing on the neurocognitive-affective gradient of the psychosis spectrum based on a well-supported conceptual model of how different conditions and symptom dimensions relate to one another (Figure 2A) (9,10,13). A schematic overview of the study is depicted in Figure 1.

**Figure 1:**
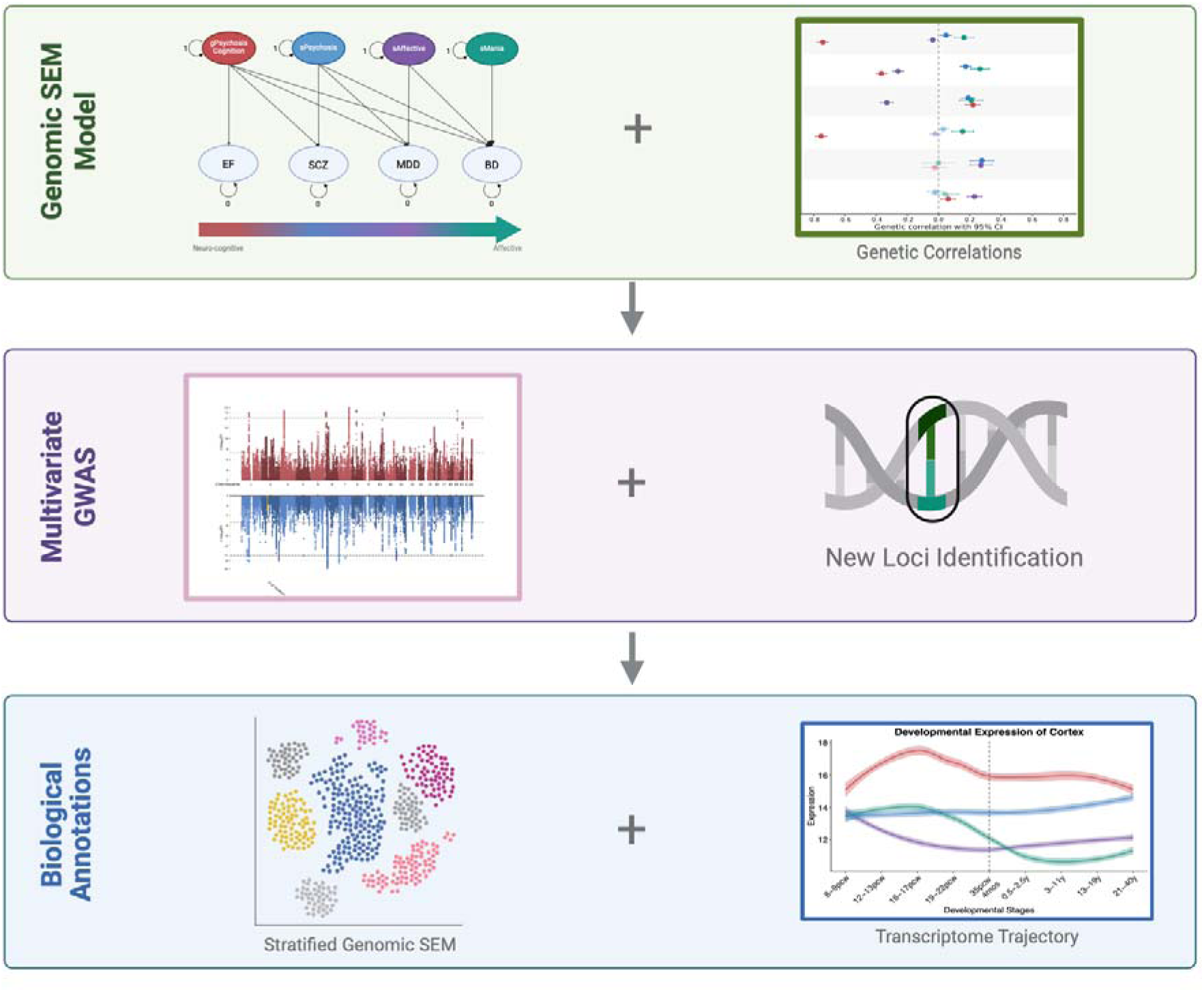
Schematic representation of the workflow. Initially, we performed a Genomic Structural Equation Model to define four latent factors representing functionally distinct portions of the psychosis spectrum. Then, we explored their genetic correlations with external traits (“Genomic SEM Model” section). Subsequently, we ran a GWAS for each latent factor and analyzed their summary statistics (“Multivariate GWAS” section). Lastly, we used Stratified Genomic SEM and transcriptome data to better understand biological characteristics of the SEM factors (“Biological Annotations” section).

**Figure 2:**
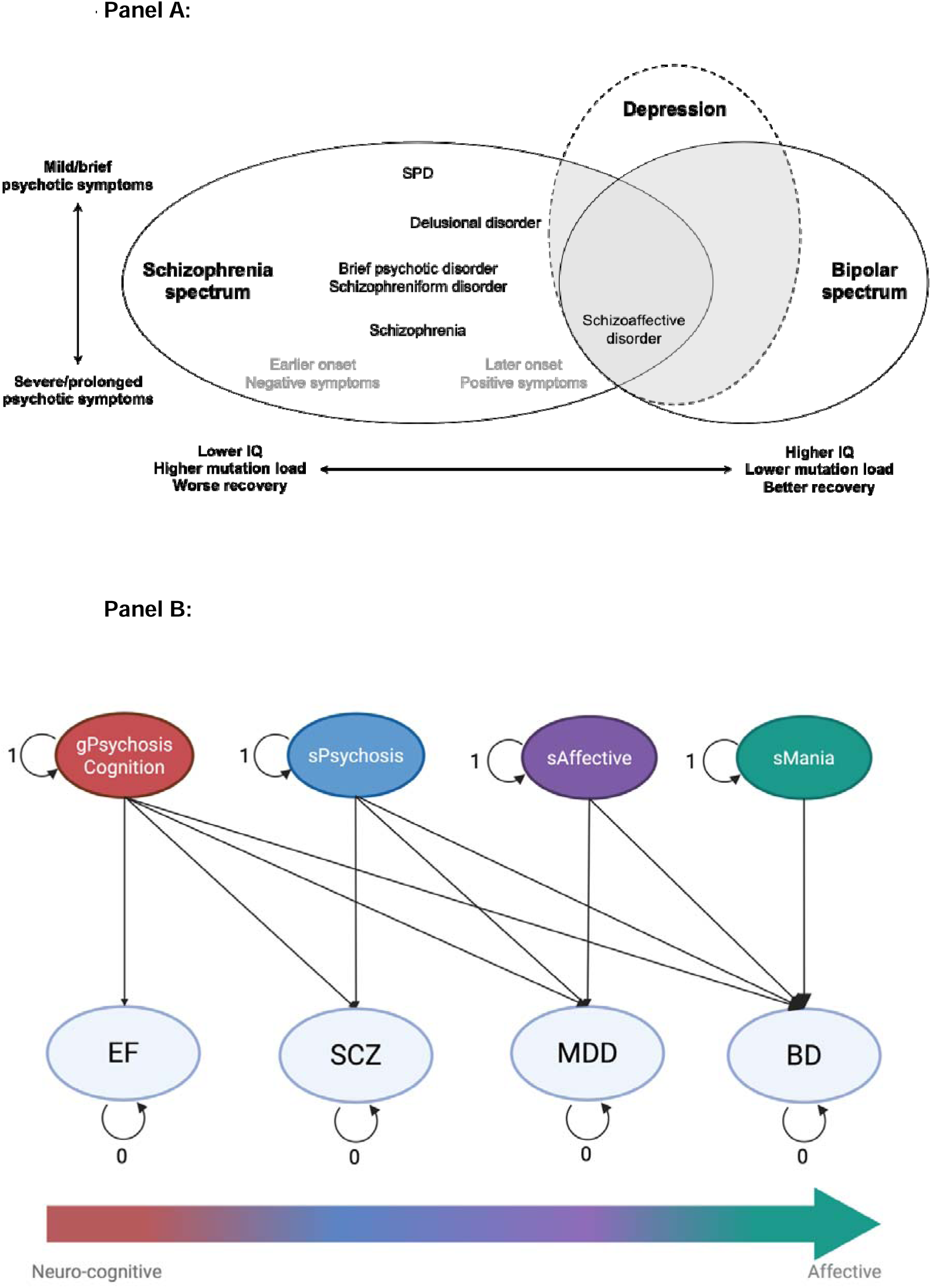
Panel A - A conceptual diagram of the psychosis spectrum. The horizontal axis represents a gradient of neurocognitive (dys)functionality. Depression is shown with a dashed contour to emphasize the fact that it only partially overlaps with the psychosis spectrum (see the main text for details). SPD = schizotypal personality disorder. Reproduced with permission from [Del Giudice 2018]. **Panel B - The Cholesky model fit in the Genomic SEM analyses**. The executive function (EF) is the leverage trait used to anchor the psychosis spectrum. The arrow highlights that the latent factors are picking portions of the gradient, blurring from the neurocognitive to the affective pole (see the main text for details). gPsychosisCognition = general factor of psychosis and cognitive dysfunction; sPsychosis = specific psychosis factor; sAffective = specific affective factor; sMania = specific mania factor; SCZ = schizophrenia; MDD = major depressive disorder; BD = bipolar disorder.

## Methods

### Theory-Based SEM Model

*Exploiting the Gradient using a SEM*. We used a SEM framework to fit a Cholesky model where diagnostic categories are ordered according to their clinical distance from a “leverage trait” that anchors the gradient. The leverage trait we chose is executive function (EF) as operationalized in Hatoum et al. (42). Executive function shows a sizable overlap with IQ and cognitive ability (42,53,54); at the same time, there are indications that it may be particularly relevant to the cognitive dysfunction in SCZ and other psychotic spectrum disorders (9,42).

Since the gradient model exploits cognition as a clear component of psychosis heterogeneity (9), we use EF as a leverage to establish the ordering of the spectrum and isolate the broad-band genetic contribution to the cognitive aspects of psychosis, thus allowing a cleaner differentiation between other sources of variation.

The ordering of indicators in the resulting Cholesky model is EF first, then SCZ, followed by MDD and lastly BD (Figure 2B). Since the role of MDD in the psychosis spectrum and, for this model, its relative position with EF, is less clear, we specified the SEM to extract a mania factor following a reasoning similar to Merola et al. (51). In this work the authors defined the mania factor as the BD component independent of MDD. However, since our objective was to partition the mood component(s) of psychosis from its neurocognitive signal, we also adjusted for SCZ and EF. As note, we did not include the SZA GWAS since it was under-powered.

In other words, this model specification leads to a good factor interpretability while respecting the well-replicated evidence of the theoretical model: SCZ shows higher cognitive impairments and represents a more neurocognitive condition than BD; while the mania component of BD represents the end point of the affective side of the psychosis spectrum (Figure 2A) (9,10,13).

The model defines four latent genomic factors: a *general factor* of psychosis and cognitive dysfunction (gPsychosisCognition); a *specific psychosis factor* independent from cognitive/executive functionality (sPsychosis); a *specific affective factor* independent of psychotic experiences (sAffective); and a *specific mania factor* independent of depression and psychosis (sMania). Together, these four factors decompose the heterogeneous genetics of psychosis into a functionally distinct set of genetic contributions.

### Genomic SEM Analyses

#### Summary Statistics

We aimed to select the most well-powered GWAS summary data from samples of European-like (EUR) ancestry. We use GWAS data from three psychosis-related disorders; Schizophrenia (55) (SCZ; Neff = 58,749), Bipolar Disorder (56) (BD; Neff = 535,720), and major depression disorder (57) (MDD; Neff = 1,152,656). Summary statistics for both BD and MDD excluded 23andMe due to these data not being publicly available. We use a GWAS of executive function (42) (EF; N = 427,037) to capture cognitive function. Although EF is often phenotypically distinct from broader cognitive function measures (42), genome-wide EF data is well-correlated with other cognitive function measures and is better-powered than other available cognitive GWAS data.

#### Fitting the Genomic SEM Model

We used Genomic SEM (58) to fit the SEM model explained above using EF, SCZ, MDD and BD summary statistics as indicators. We started the analyses munging the summary statistics: we removed SNPs with imputation score (INFO) < 0.9, Minor Allele Frequency (MAF) < 0.01, and those that were not present on the HapMap3 reference panel. Then, we used the munged summary statistics as input for Genomic SEM. First, the ldsc() function was used to create the variance-covariance matrix (*S*) and its sampling variance matrix (*V*). The diagonal of the S matrix contains the SNP-based heritability for each trait; its off-diagonal elements, instead, are populated by all the pairwise co-heritability (*Supplementary Fig. 1*). The diagonal elements of the *V* matrix are the squared standard errors of the diagonal elements of *S*; the off-diagonal elements, instead, are the pairwise covariances of the pairwise co-heritabilities in *S*. We used both the *S* and the *V* matrices to fit the SEM model, where the elements of the *V* matrix are used as weights of the Diagonally Weighted Least Square (DWLS) convergence algorithm.

#### Model Specification and Factor Coding

All the factors in the Cholesky decomposition are identified using unit-variance scaling. To achieve a better interpretability, we decided to reverse-code the gPsychosisCognition factor, leading to a negative unit loading on the EF indicator and positive loadings on SCZ, MDD, BD (Figure 3). In this manner, the gPsychosisCognition factor can be interpreted as a general psychosis dimension with a cognitive impairment.

**Figure 3:**
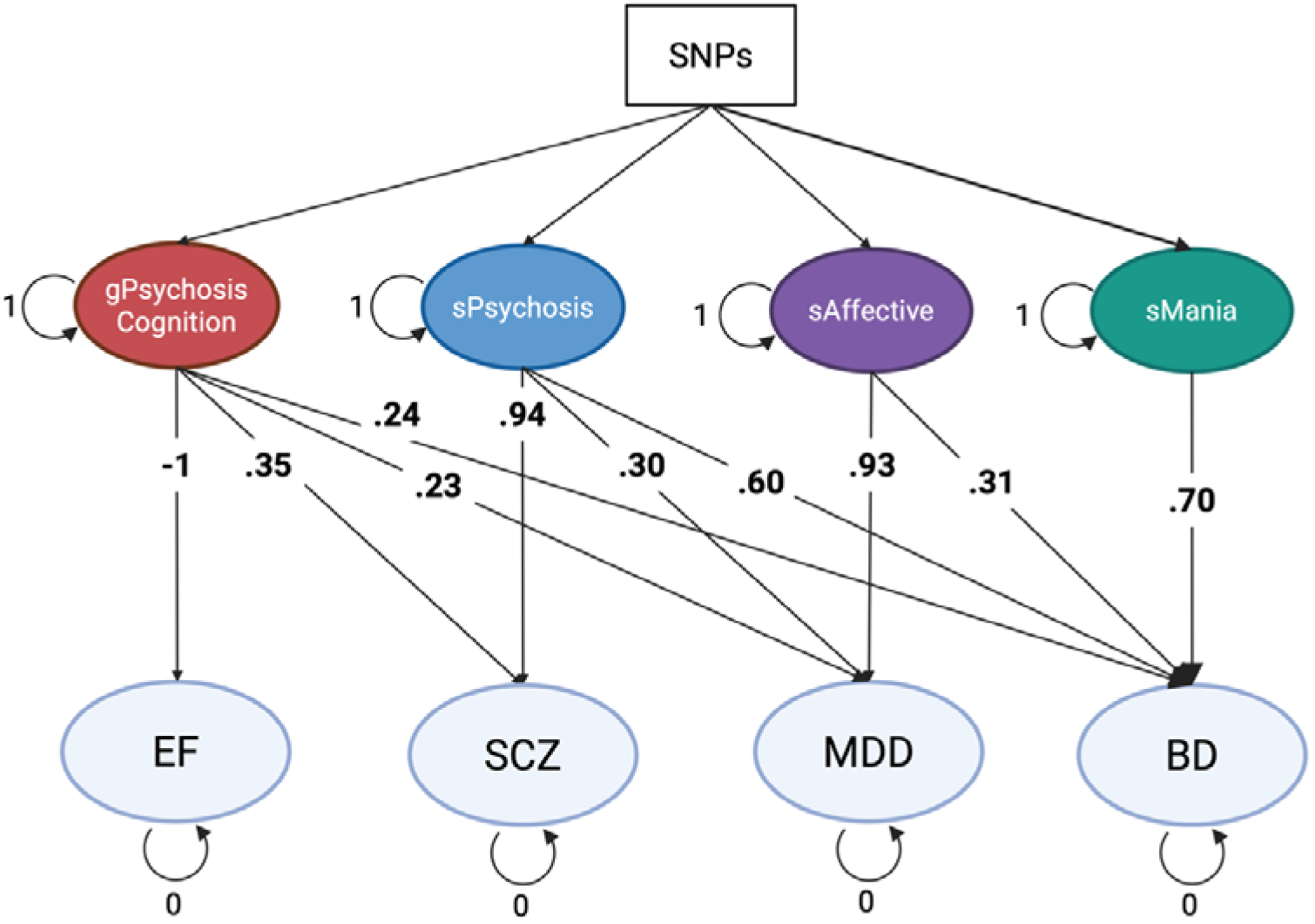
The Genomic SEM model fit with SNPs regressed on each factor. Bolded loadings are significant at *p <.*001. The SNPs regression allows the identification of one multivariate GWAS for each Cholesky factor.

#### Stratified Genomic SEM

We used stratified Genomic SEM (s-Genomic SEM) (11) to partition the variance of the Cholesky factors into functional annotations. The s-Genomic SEM is a two-step analysis: we firstly created the zero-order matrices for the annotations and then used the enrich() function to partition the heritability of the Cholesky factors within annotations. We specified two sets of annotation: the Baseline V2.2 and the “superclusters” from Siletti et al. (59). The latter set is based on single-nucleus RNAseq data, allowing us to better understand cell types and topological enrichments of our factors. We used only the binary annotations and removed the ones with a pre-post-smoothing difference > 1 (methods details in *Supplementary Materials*).

### Multivariate GWAS Via Genomic SEM

#### Multivariate GWAS

Two steps are required to perform a multivariate GWAS on the latent factors. We firstly used the sumstats() function to standardize the individual summary statistics, so SNP alleles were aligned in the same direction and SNP effect sizes were scaled using the EUR ancestry 1000 Genome Phase 3 reference panel. Subsequently, we used the userGWAS() function to compute a regression of the SNPs on the latent factors: SNPs are added in the *S* and *V* matrices, and regression paths are estimate for each SNP on the four latent factors. Once the multivariate GWAS summary statistics are extracted, the sample size estimation (N^”^) was calculated using the function introduced in Demange et al. (60) after applying a MAF filter that requires SNPs to have MAF ranging from 10%-40% (details in *Supplementary Materials*).

#### Novel SNPs

Since multivariate GWAS techniques increase discovery power, we checked whether any novel SNPs were identified. We defined novel SNPs as SNPs that were not previously associated with the SEM indicators or with any other trait in the GWAS catalog. This was done in two steps: we first checked for “novel SNPs” among the Cholesky indicators using the get_novel() function implemented in GWASLab [GWASlab preprint]. Since the function requires fetching traits from *GWAS Catalog*, we queried “EF measurement” (EFO_0009332), BD (MONDO_0004985), SCZ (MONDO_0005090), and MDD (MONDO_0002009). Subsequently, we used the get_associations() function to check if the “novel SNPs” were significant in traits not included in the first analysis. SNPs that showed no association in both steps were labeled as “novel SNPs”.

### Genetic Correlations

#### Correlations with other Traits

We used Genomic SEM to analyze the genetic correlations between the Cholesky factors and different external traits. We selected eight clinical, cognitive, personality, and life-history traits with available GWAS (selection reasoning details in *Supplementary Materials*): Autism spectrum disorder (ASD) (61), Neuroticism and Openness (62), Educational Attainment (EA) (63), IQ (64), Cog and NonCog components of EA (60) and Number of children Ever Born (NEB) (65). Of note, we used NEB as a rough proxy of evolutionary fitness to explore the possibility that different parts of psychosis could have opposite pattern of association with NEB. This was motivated by evolutionary models that posit how psychosis could have persisted due to direct or indirect evolutionary adaptiveness of this trait (10).

The same pipeline used for the Cholesky model was used to munge the external traits and to create the *S* and *V* matrices. To test correlations between the Cholesky factors and the external traits, we iteratively performed a Genomic SEM model for each external trait in which we built the Cholesky decomposition while letting the external trait correlate with the latent factors. To account for multiple testing an FDR correction was applied across all the combinations of factor and external trait.

We used the delta method to test if two different Cholesky factors showed statistically different correlation with the same external trait (e.g. *rg_F1—traitŽ_* vs. *rg_F2—traitŽ_*). For each pair of factors that showed significant correlation with the external trait, we set a function *g:— rg_F1—traitŽ_ — rg_F2—traitŽ_* and tested it against the null hypothesis *H_0_: g = 0* (i.e. constraining the correlations to be equal), extracting the p-value conditioned on this hypothesis. The delta method computes the p-values defining the *Vax(g) = JV_θ_J^T^*, where *J* is the Jacobian vector of the parameters θ of the model-implied matrix (θ), and *V_θ_* is the variance-covariance matrix of the estimated parameters θ. Unlike the likelihood ratio test, using the delta method through the g function allowed us to directly test differences across coefficients.

### Lead Loci, Gene Mapping and Transcriptome Trajectories

#### Lead Loci

We analyzed our multivariate GWASs using FUMA v1.5.2 (66) to find independent and lead SNPs. Among the SNPs that reached the significance threshold (*p* < 5*e* — 8) we set the threshold for “independent SNPs” as *r^2^* ≥ 0.6 and a secondary threshold for “lead SNPs” as *r^2^* ≥ 0.1. To identify independent genetic risk loci, we used the Phase3 1000 Genome reference panel for the EUR ancestry with a window of 250kb.

#### Gene Mapping

To understand the genes implicated by the multivariate GWASes, we used hMAGMA (v1.10)(67) that uses both positional mapping and regulatory effects (from chromatin interactions Hi-C and eQTL) to test the effect of the SNPs on the genes. We used both adult and fetal annotations for pre-and post-natal human cortex. Only Bonferroni significant protein-coding genes located outside of the Major Histocompatibility Complex (MHC) were selected. We excluded the MHC due the complex LD structure of this region. We chose Bonferroni correction to provide a more stringent correction favoring stronger biological signal, aiming to obtain more biologically meaningful downstream analyses. The MAGMA gene-set analysis was performed using the SNP2GENE step in FUMA (66) using default settings.

#### Transcriptome trajectories

We used data from PsychENCODE (68) to analyze the expression pattern of the Cholesky factors across the lifespan. This database provides RNA-seq data from post-mortem samples that cover approximately all the life stages, from pre-natal to adulthood. PsychENCODE data underwent an extensive pipeline of quality checks and a RPKM normalization that accounts for both sequencing depth and gene length (see Li et al. (68) for details). The transcriptomic trends are computed averaging the expression of all the factor specific genes extracted with hMAGMA. Since expression in the brain is both temporally and spatially differentiated, we separately averaged across the whole cortex, frontal lobe, prefrontal cortex and striatum to better appreciate regional expression differences throughout the lifespan. For the prenatal samples, we used significant genes from hMAGMA annotated with fetal annotations, and adult annotations were used to select genes in postnatal samples. A local regression (LOESS) was used to plot the transcriptome trajectories.

Claude was used to troubleshoot codes to resolve coding errors.

## Results

### Genomic Dissection of the Psychosis Spectrum

The Genomic SEM model revealed that the four factors independently contributed genetic variance to the psychosis spectrum, with all the loadings significant (*p <.*001) (Figure 3, Table 1). Since the model is just-identified, fit indices are not available. The gPsychosisCognition factor loadings were stronger for SCZ (. 35) than MDD (. 23) and BD (. 24); moreover, BD and MDD loadings were similar, suggesting similar involvement of these clinical conditions with EF. These observations are in line with Hatoum et al. (42)(Fig. 5A). The sPsychosis factor loadings were. 94 for SCZ,. 30 for MDD, and. 60 for BD, indicating that the genetic overlap between MDD and psychosis is driven not only by cognition, but also by additional non-cognitive shared components. The sAffective factor, as expected, showed a strong loading with MDD (. 93) and a smaller but significant loading with BD (. 31), capturing the mood component of BD. The sMania factor replicated the findings by Merola et al. (51), identifying a BD-specific genetic component independent from MDD (Figure 3, Table 1). The estimated N” for the factors were 410,883 for gPsychosisCognition; 78,475 for sPsychosis; 680,152 for sAffective and 114,849 for sMania.

**Table 1.**
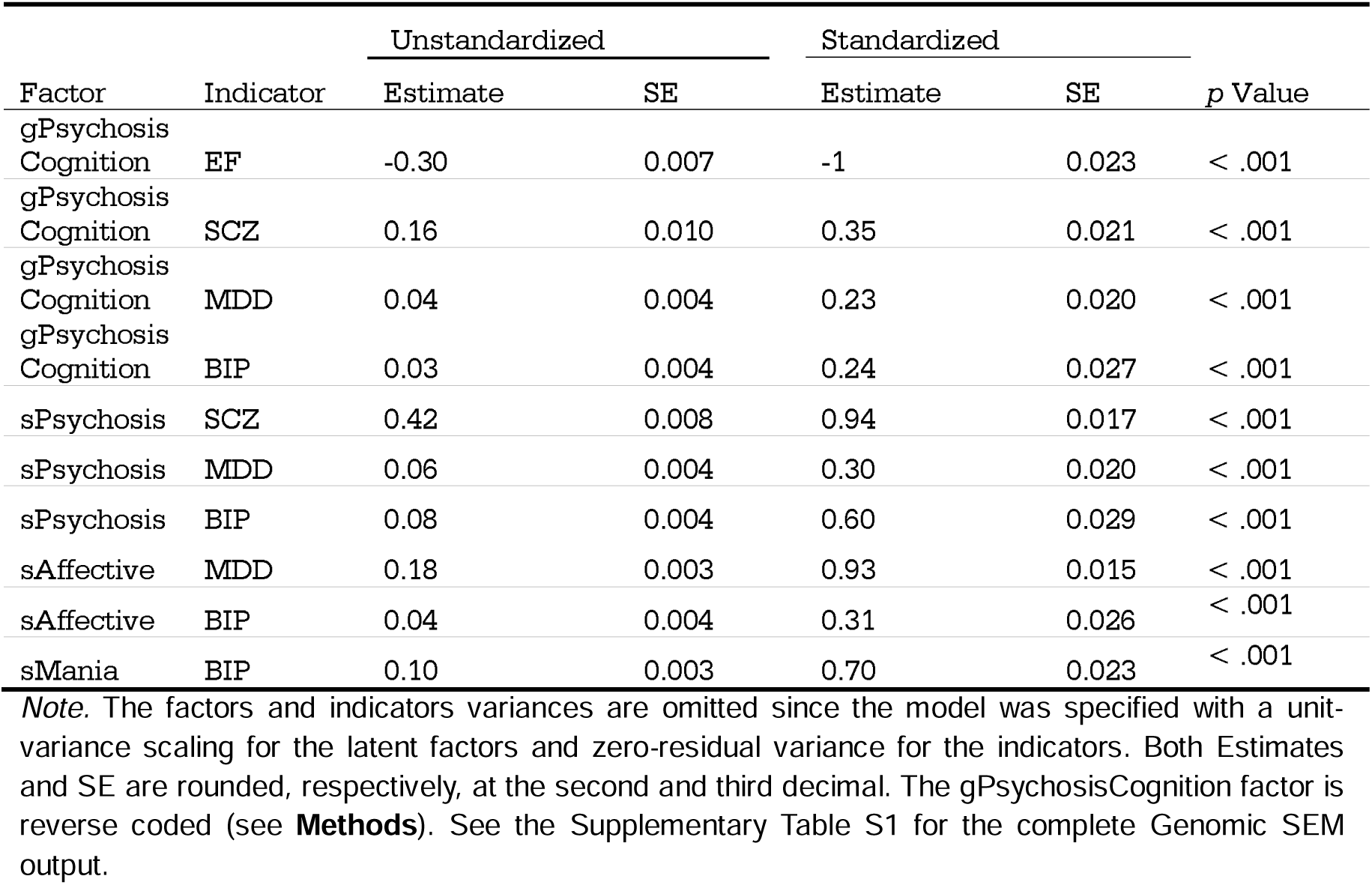
Genomic SEM results of the Cholesky model.

The FUMA analysis revealed 95 risk loci for gPsychosisCognition, 106 for sPsychosis, 134 for sAffective and 23 for sMania. We identified one new risk locus for sPsychosis (rs6721531, 2p12), three for sMania (rs833638, 3p14.2; rs146786560, 11q23.3; rs10131905, 14q23.2) and, finally, sAffective showed 10 new risk loci (Supplementary Table S2, *Supplementary Fig. 2 and 3*). We did not find any novel loci for gPsychosisCognition.

### Correlations with Other Traits

*Genetic Correlations*. The associations among the factors and external traits revealed distinct relationships (Figure 4, Supplementary Table S3). gPsychosisCognition showed strong negative correlations with Intelligence (*r_g_* = —.74, *SE* =.01) and EA (*r_g_* = —.36, *SE* =.01), while a positive correlation with NonCog (*r_g_* =.22, *SE* =.02). Moreover, gPsychosisCognition exhibited a positive genetic correlation with Neuroticism (*r_g_*=.26, *SE* =.02) while a non-significant genetic correlation with Openness. The negative correlations between EA and Intelligence and the gPsychosisCognition factor were consistent with gPsychosisCognition capturing cognitive impairment, which is aligned with the factor coding (**Methods**).

**Figure 4:**
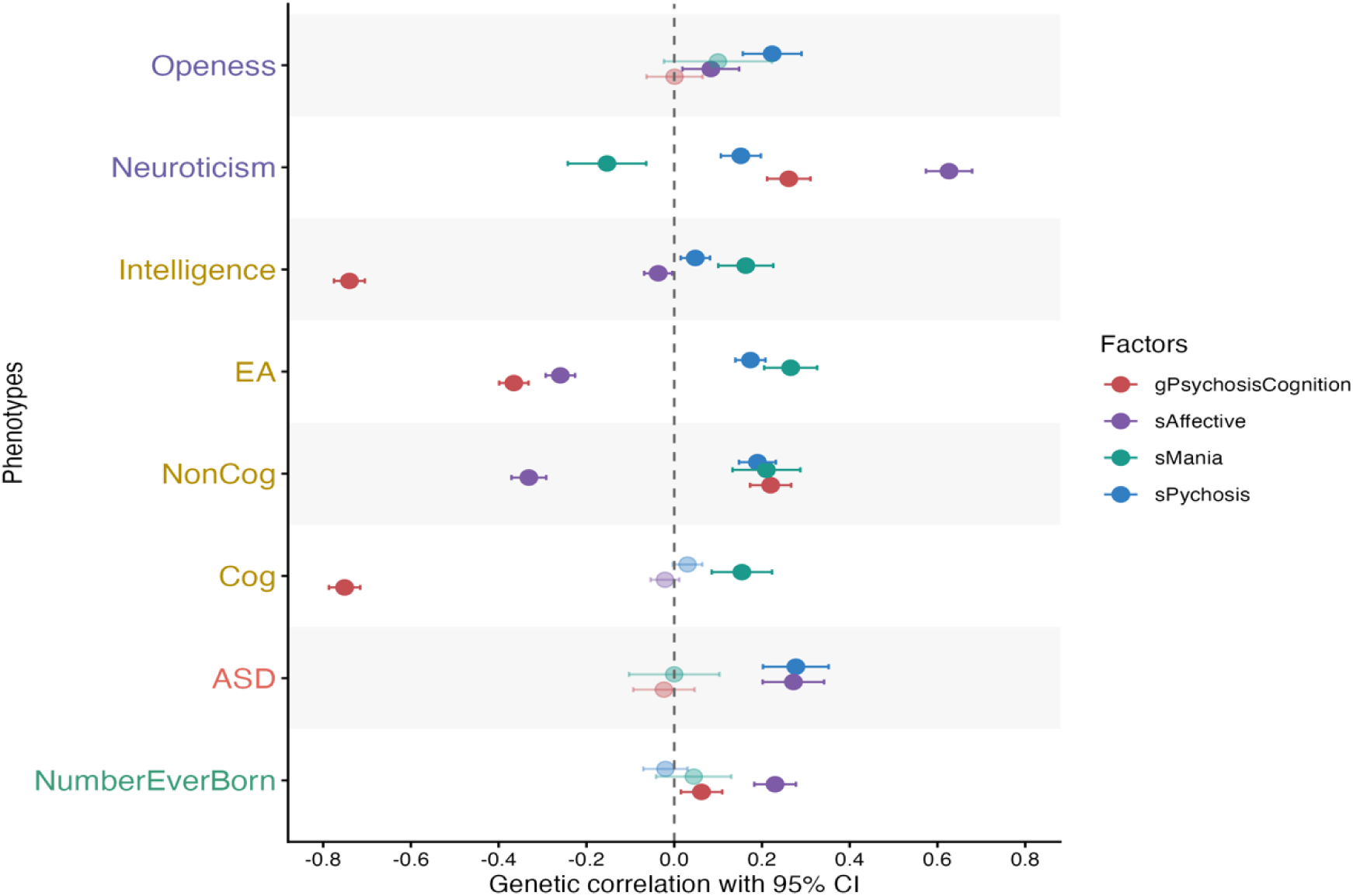
Genetic correlations between the Genomic SEM factors and external traits. The x-axis represents the genetic correlation with 95% CI. The points with low opacity are non-significant, whereas the opaque points are significant. The external phenotypes are grouped by category, where purple are the personality traits, the yellow are the cognitive traits (EA = Educational Attainment, NonCog = the non-cognitive component of EA, Cog = the cognitive component of EA); in red is Autism Spectrum Disorder (ASD) and green the Number of Children Ever Born (NumberEverBorn) that represents a proxy for evolutionary fitness.

**Figure 5:**
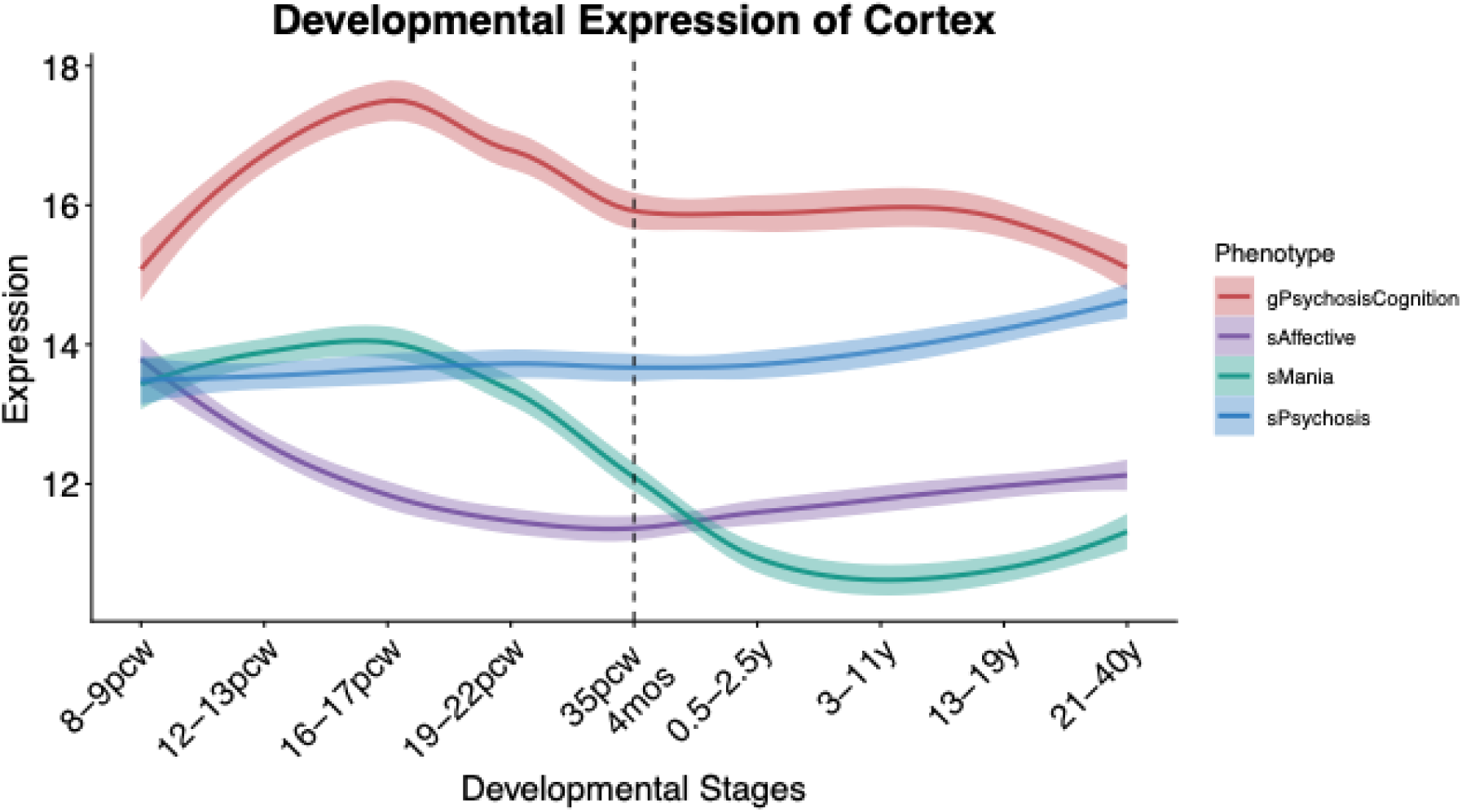
Whole-cortex transcriptome trajectories across development. The x-axis shows the developmental stages with the vertical dashed line representing time at birth. pcw = post conception weeks, mos = months and y = years. The y-axis represents the normalized expression averaged across the Bonferroni significant genes from the hMAGMA analysis.

sPsychosis showed positive genetic correlations with Openness and Neuroticism (*r_g_* =.22, SE =.03; *r_g_* =.15, SE =.02) along with EA (*r_g_* =.17, SE =.01), NonCog (*r_g_* =.19, SE =.02) and ASD (*r_g_* =.27, SE =.03).

sAffective showed positive genetic correlations with Neuroticism (*r_g_* =.62, *SE* =.02) ASD (*r_g_* =.27, SE =.03) and NEB (*r_g_* =.22, *SE* =.02), while negative genetic correlations with EA and NonCog (respectively, *r_g_* = —.25, *SE* =.01; *r_g_* = —.33, *SE* =.02).

Finally, sMania showed positive genetic correlations with Intelligence, EA and NonCog (respectively, *r_g_* =.16, *SE* =.03; *r_g_* =.26, *SE* =.03; *r_g_* =.21, *SE* =.04) while negative with Neuroticism (*r_g_* = —.15, *SE* =.04). This is in line with Merola et al., 2026 that identified a negative genetic correlation between the mania factor and Neuroticism.

The delta method revealed that most of the genetic correlations were pairwise statistically different. Although, there were some exceptions: the correlations of sPsychosis and sAffective with ASD were not statistically different (*p* =.92) along with the correlations of sPsychosis and sMania with EA (*p* =.09). Neither sPsychosis and sAffective (*p* =.08), nor sPsychosis and sMania (*p* =.10) showed statistically significant differences in correlations with Intelligence. For the NonCog component, none of the pairwise tests between gPsychosisCognition, sPsychosis and sMania were statistically significant (*p* >.05) (Supplementary Table S4).

### Gene Mapping and Transcriptome Trajectories

To better understand the biological characteristics of the Cholesky factors, we first mapped the SNPs onto genes using hMAGMA, identifying 275, 290, 53 and 224 Bonferroni significant genes for gPsychosisCognition, sPsychosis, sMania and sAffective (Supplementary Table S5).

Subsequently, we used the significant genes to better understand the temporal trends of cortex expression across lifespan. The trajectories (Figure 5) revealed that the gPsychosisCognition factor had a prenatal peak, sPsychosis demonstrated a stable pattern of expression, while sAffective and sMania showed a prenatal peak with a later descending trajectory. This is in line with previous findings showing that SCZ genes are both pre-and postnatally expressed (69–72) while BD is more prenatally expressed (71). We also separately analyzed the prefrontal cortex (PFC), dorso-lateral prefrontal cortex (DFC), and the striatum, as they are often impaired in psychosis (73–75). Although the trends remained mostly unchanged for PFC and DFC, striatum trends showed some degree of specificity: sMania and sPsychosis showed the same time trend with a peak at the 19-22 post conception week window; a mostly constant expression for sPsychosis and a mostly postnatal peak for gPsychosisCognition. Overall, we did not detect a strong single-area specificity, especially for the cortical subregions (*Supplementary Fig. 4 to 6-Supplementary Tables S10 to S13)*.

**Figure 6:**
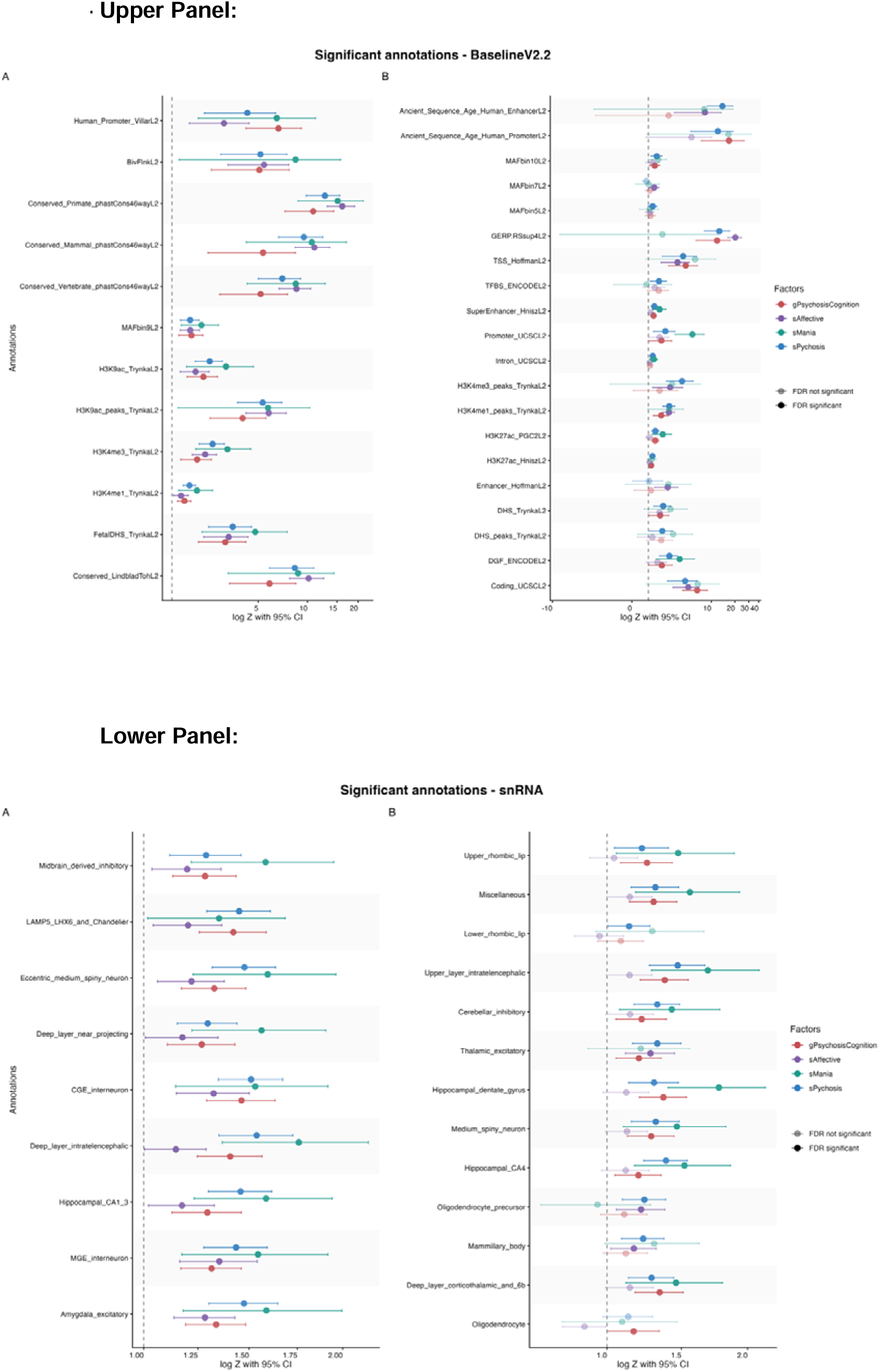
Significant annotations from s-Genomic SEM analysis. The upper panel contains the results from annotations in the BaselineV2.2 model while the lower panel contains results from annotations based on the snRNA data from [Siletti et al., 2023]. For each panel, the A plots contain the annotations that are significant for all the factors, whereas the B plots depict the annotations that exhibit distinct patterns of significance, with at least one factor significant and at least one factor non-significant. The x-axes represent the log10 of the enrichment Z-score with 95% CI. The y-axes represent the respective annotations. The opaque points and whiskers are FDR significant, the non-fully opaque are non-FDR significant.

### Biological Annotations with s-Genomic SEM

The BaselineV2.2 (Figure 6) annotations revealed similar enrichment across factors for evolutionary conserved annotations (Conserved primate *p* <.001, mammal *p* <.01 and vertebrate *p* <.004), expression regulation in histone 3 and FetalDHS_Trynkal (*p* <.01) (Supplementary *Tables S6 to S9*), highlighting both the key importance of chromatin organization and early developmental processes for the psychosis liability (76,77). Moreover, factors showed different enrichments pattern across regulatory processes (Promoter_UCSCL2: p >.05 only for sAffective, Intron_UCSCL2: *p* >.05 for sAffective and gPsychosisCognition, TFBS_ENCODEL2: *p* <.05 only for sPsychosis, TSS_HoffmanL2: *p* >.05 for sMania). Although these results represented biological specificities between factors, they underlined the role of non-coding regions in mental health etiology (78). Moreover, the sMania was the only factor not enriched for the coding regions (Coding_UCSCL2 *p =*.053); this was replicated also in FUMA analyses where exonic regions were not enriched.

The single-nucleus enrichments show both similarities and differences between factors (Figure 6). Both Deep-layer (p < 0.05) and Upper-Layer Intratelencephalic enrichments were significant (p ś.004), with the exception for sAffective in the Upper-Layer (*p =*.057), strengthening the importance of these cell types in SCZ (79) and, more generally, as a broad psychosis liability encompassing both clinical and cognitive traits (see fig. 2A Yao et al. (80)). The enrichment highlights the role of amygdala (Amygdala excitatory, *p <.*01) and hippocampal regions, where CA1-3 (p <.03) were significant for all the factors but CA4 (sAffective: *p =*.1) and dentate gyrus (sAffective: *p =*.08) were not significant for sAffective. In line with Yao et al. (80), non-neuronal clusters tended to not be involved in psychosis (Astrocytes p >.22, Microglia p >.15, Bergmann glia p >.09, committed oligodendrocytes precursors p >.15); however, we identified some exceptions: oligodendrocytes were significant for gPsychosisCognition (*p =*.04) and oligodendrocyte precursor were significant for sPsychosis (*p <.*001) and sAffective (*p =*.01). Moreover, both interneurons (CGE_interneurons p ś.01, MGE_interneurons *p <.*01) and medium spiny neurons (Eccentric_medium_spiny_neurons *p <.*01, Medium_spiny_neurons non-significant only for sAffective *p =*.06) were significant, underlying the relevance of GABAergic processes in the psychosis etiology (77,79).

## Discussion

We combined Genomic SEM with a clinically grounded conceptual model to partition the psychosis spectrum into functionally distinct sources of genetic liability. We identified 18 loci that were not previously associated with the psychosis domain. Moreover, the GWASLab analyses revealed that four of the new loci were associated with physical and medical traits (asthma, serum lead level, metabolic syndrome and chronotype – Supplementary *Table S2*), replicating previous demonstrations of genetic overlap between mental and physical illness (81). We replicated the association between respiratory and metabolic functionality with thought disorder (81), along with the relationship between lead exposure and SCZ (82).

The genetic factors generally replicated phenotypic associations among the psychosis spectrum and cognition (Figure 2A and 2B); for example, sMania shows a stronger positive correlation than sPsychosis and sAffective with Intelligence while, as expected, gPsychosisCognition represent the strongest negative correlation with cognitive ability. In other words, as one moves towards the affective pole, factors increasingly show positive genetic correlations with Intelligence, thus replicating the pattern observed at the phenotypic level (9,13). However, the NonCog component of EA did not follow the same pattern and instead showed a specific negative correlation only with the sAffective factor. In particular, the correlations of gPsychosisCognition and sAffective with Cog vs. NonCog highlighted opposite patterns: gPsychosisCognition showed substantial negative genetic correlations with Cog, but positive correlations with NonCog. On the other hand, sAffective showed weak, and mostly non-significant, genetic correlations with Cog but a strong negative correlation with NonCog. These patterns are aligned with previous literature that report a complex interplay between psychosis and academic success (44,83), where the different cognitive components play an opposite role on psychosis (60,84).

Openness showed the strongest correlation with the sPsychosis (non-cognitive psychosis) factor pointing to a potential functional aspect of psychosis proneness. Previous literature demonstrated that the positive relationship between Openness, SCZ and cognition is driven by higher-order cognitive functions (60,84) that might drive the associations with creativity (85–87). These patterns has been also observed in psychology literature where the enhanced creative thinking in psychosis is accounted by Openness (88–90) that, in turn, is more associated with crystallized intelligence (91). However, additional work on the interplay between cognition, different aspects of psychosis liability and creativity are needed to better understand their relationship.

We also replicated results from Merola et al. (51) regarding the positive genetic correlations of mania with EA and NonCog. Moreover, the negative associations of mania with Neuroticism (51) suggest that it could represent a specific liability rather than a predisposition to multiple psychopathological domains(1,92). In contrast to some previous theories (e.g. [Craddock & Owen] (13)), the genetic associations between psychosis and ASD were not accounted for by the general cognitive factor (gPsychosisCognition), but by non-cognitive components (sPsychosis and sAffective). However, the intersection between low intelligence, ASD and psychosis might be due mainly to rare variants that were not considered in this study (13,93). Notably, the involvement of sAffective with ASD is in line with previous literature that has highlighted the role of mood regulation and co-occurrence of internalizing psychopathology in ASD (61,94,95). Future research might explore the overlap between different aspects of psychosis proneness and specific dimensions of the autism spectrum.

The dissection into functionally distinct components also allowed us to investigate the so-called “evolutionary paradox” of psychosis: why do harmful and highly heritable conditions persist at relatively high incidences, instead of being kept to very low frequencies by selection? In our data, the fitness proxy represented by offspring number (NEB) was positively correlated with both gPsychosisCognition and sAffective. The positive correlation between gPsychosisCognition and NEB may be driven by the relationship between EA and lower fertility (96). Of note, due to the factor coding, higher scores on gPsychosisCognition imply lower cognition (**Methods**), in line with the expected negative genetic correlation between EA and NEB (96).

All the factors were enriched for the evolutionary conserved regions, while the sMania was consistently non enriched for both ancient annotations and GERP. Overall, these patterns replicate the negative selection found for complex traits (86,97) but suggest weaker evolutionary pressures for sMania.

We replicated well-known biological associations, such as the role of GABAergic neurons in psychosis (79), the higher involvement of neuronal vs. non-neuronal cells, and the contributions of different cortical layers (79,80). Moreover, we were able to better characterize the processes uniquely involved in our factors (Figure 6B): sAffective was the only factor with a (mostly) non-significant enrichment for hippocampus and cerebellum, thalamic excitatory neurons were non-significant only for sMania, while oligodendrocytes were uniquely involved in gPsychosisCognition. Finally, we observed that the factors exhibited non-trivial overlap across enrichments (Figure 6A).

The transcriptomic trajectories revealed different patterns across the lifespan, where factors showed differing transcriptomic trajectories. This result extends the notion that psychosis-related conditions are composed by genes that show remarkable temporal pattern differentiation (71,72) involving both pre-post-natal processes and remarking its (partial) neurodevelopmental origin. Moreover, the gPsychosisCognition and sAffective prenatal peak is in line with previous evidence that found a prevalent prenatal expression for Intelligence and Neuroticism (98). Moreover, the limited spatial differentiation is in line with previous works (98,99) that highlighted a prevalence of temporal differentiation over regional specificities, driving both normative cortical development and disease similarities (Fig. 2b Liu & Shimogori (98)). These evidence stress the importance of temporal regulation in psychosis, revealing sensitive developmental stages implicated in the disease etiology along with information about the phenotypical presentation (98).

### Limitations

There are some limitations of note. First, the Genomic SEM factors did not account for the same amount of variance, leading to differences in power that could be roughly captured by the N^”^as well as by the mean y^2^. Power differences could have hindered our ability to detect true biological processes that uniquely characterize each factor, leading to Type II errors due to low statistical power. Moreover, the model is based on GWAS studies conducted on participants of European-like ancestry, limiting the generalizability of our results to other populations. Additionally, the limited resolution of the annotations specified in the s-Genomic SEM hindered the possibility to find specific biological processes between the factors. Lastly, the transcriptome trajectories assume a homogeneous temporal trend across genes of the same factor. Moreover, they were modeled non-parametrically, limiting our ability to build statistical tests on trend shapes.

## Conclusion

This work highlights the complexity of the psychosis spectrum, as its genetic components show differential associations with both functional (e.g. Openness, creativity, etc.) and non-functional (e.g. cognitive impairment) traits, consistent with the idea that a multicomponent genetic architecture underlies the phenotypic heterogeneity of the psychosis spectrum. We discovered new genetic loci associated with psychosis domain and identified factor specific biological mechanisms that, in turn, suggested more nuanced biological implications.

## Data Availability

All data produced in the present study are available upon reasonable request to the authors

## Acknowledgements

This work used the High Throughput Computing Facility at the Center for Genome Sciences and Systems Biology at Washington University in St. Louis.

