## Supplementary Methods for "Parsing the Functional Heterogeneity of the Psychosis Spectrum: A Clinically Informed Application of Genomic Structural Equation Modeling"

### Supplementary Information

##### Stratified Genomic SEM

Stratified Genomic SEM (s-Genomic SEM) (1) is based on a multivariable extension of stratified LDSC (s-LDSC) (2), which partitions the heritability into annotations. We define annotations as groups of genes that share common characteristics, such as tissue expression, cell-type, evolutionary history, biological processes and more. The s-Genomic SEM expands the s-LDSC technique into the SEM framework, enabling us to test enrichment for the parameters of the model-implied matrix. The s-Genomic SEM pipeline is constituted of two steps, each performed using a function directly implemented in the Genomic SEM package.

The first step uses the s_ldsc() function to create different matrices objects for each annotation: a zero-order genetic variance-covariance matrix and a sampling covariance matrix. In other words, this function creates the s-LDSC objects based on the specified annotations that will be used in the next step.

The second step takes the matrices defined by the s_ldsc() function and performs the actual enrichment computation. In particular, the enrich() function uses a reference panel (here, 1000Genome for European ancestry) to add LD weights and SNPs frequencies. Subsequently, it estimates the enrichment for the selected annotations. We used only zero-order matrices for the binary annotations and removed the ones with a pre- post- smoothing difference $>1$, as suggested by the Genomic SEM authors. We applied FDR correction for enrichment p-values to control for multiple testing.

##### Factors $\hat{N}$ Estimate from Multivariate GWAS

To estimate the $\hat{N}$ of each factor’s summary statistics we used the Demange et al. (3) pipeline. After selecting SNPs with MAF ranging from 10% to 40%, we applied the formula proposed by the authors. It solves for per-SNP sample size $n_{i}$ using the relationship between the estimated effect size ${est}_{i}$, Z-score $Z_{i}$ and the variance for SNP $i$ defined as $\sigma_{i}^{2}=2* {MAF}_{i}*\left( 1-{MAF}_{i} \right)$. A lambda correction factor is applied that represents the residual heritability of each factor.

$$n_{i}= \frac{{(Z_{i}/({est}_{i}* \lambda))}^{2}}{\sigma_{i}^{2}}$$

For each latent factor $F$, the formula finds $\hat{N_{F}}$ by averaging over $n_{i}$.

$$\hat{N_{F}}= E(n_{i})$$

##### Correlation Traits Selection

We decided to better characterize the Genomic SEM factors using genetic correlations with external traits. We selected these traits based on their relevance to the psychosis domain and on their relatedness with open scientific questions.

In particular, SCZ shows some phenotypic and genetic overlap with ASD (4–6), but their relationship is not well understood and there are still a number of competing models (6–8). With regard to personality factors, Openness has been repeatedly linked to psychosis (9–11), and Neuroticism has been shown to be a strong transdiagnostic predictor of psychopathology, particularly (but not limited to) the internalizing spectrum (9,12). In addition, the relationship between IQ and EA with psychosis seems to be driven by a multicomponent mechanism that scholars are working to shed light on (13,14). Finally, we used NEB as a rough proxy of evolutionary fitness to explore the possibility that different components of the psychosis gradient show divergent associations with NEB patterns. This was motivated by the idea that psychosis has not been wiped out by natural selection due to direct or by-product evolutionary advantage associated with some of the psychosis-related traits and their underlying genetic components (15).

15.	Del Giudice M. Evolutionary psychopathology: A unified approach. Oxford University Press; 2018.
