## Supplementary Figures for "Parsing the Functional Heterogeneity of the Psychosis Spectrum: A Clinically Informed Application of Genomic Structural Equation Modeling"

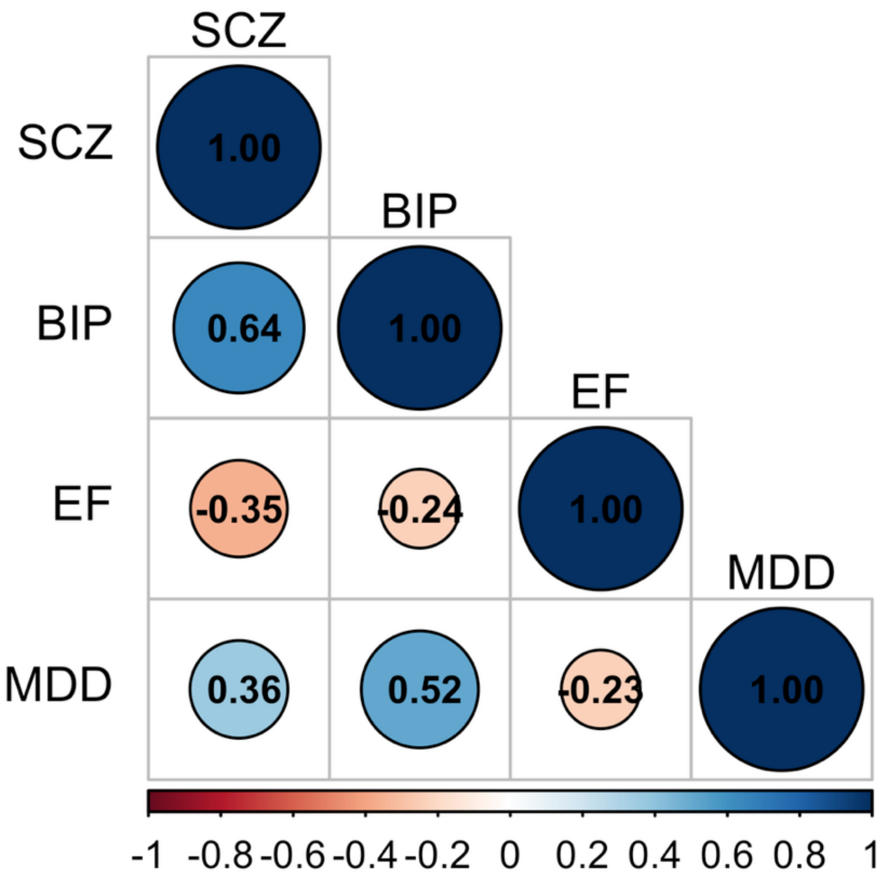


***Supp Fig.1:*** **Genetic correlation matrix among the indicators of the Cholesky model.** This matrix was obtained standardizing the variances-covariances of the $S$ matrix with the heritability of the trait(s). SCZ = schizophrenia; MDD = major depressive disorder; BD = bipolar disorder, EF = executive functions.

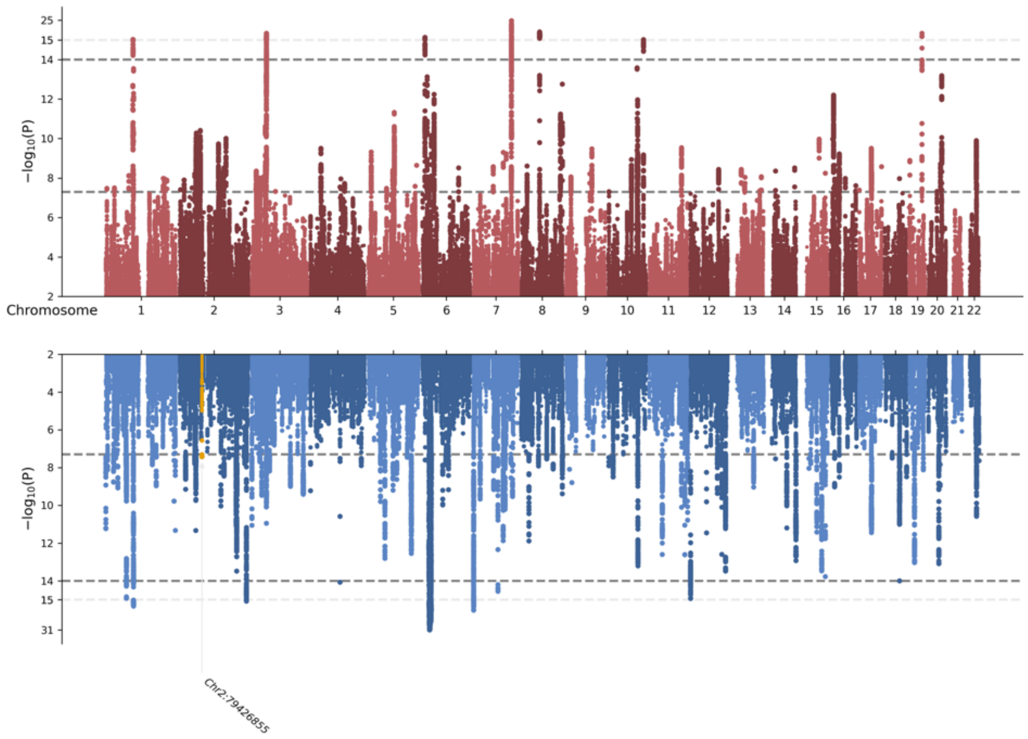


***Supp Fig.2:*** **Multivariate GWAS gPsychosisCognition and sPsychosis**. Miamiplot for gPsychosisCognition (in red) and sPsychosis (in blue). The novel SNPs in each factor are annotated with their CHR:BP position and are colored in yellow.

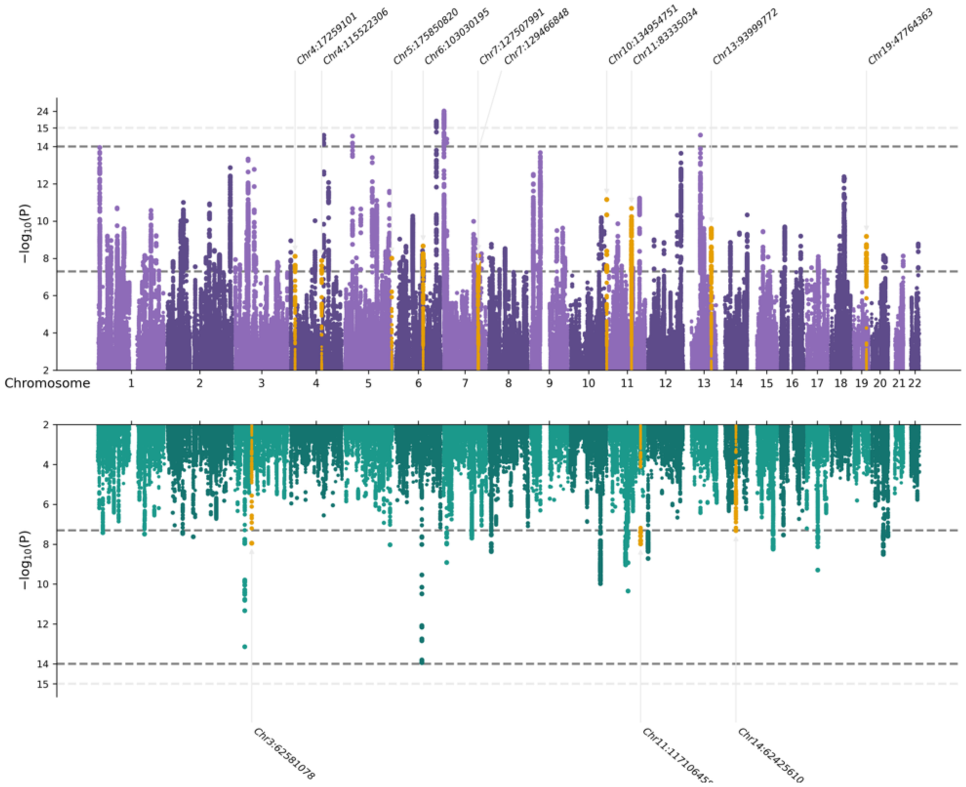


***Supp Fig.3:*** **Multivariate GWAS for sAffective and sMania**. Miamiplot for sAffective (in purple) and sMania (in green). The novel SNPs in each factor are annotated with their CHR:BP position and are colored in yellow.

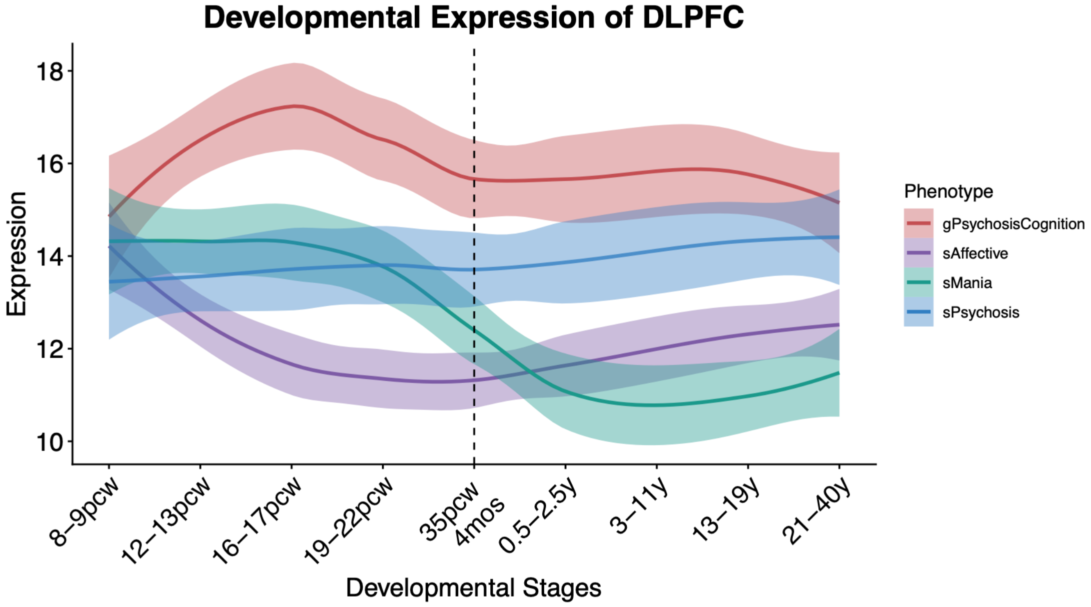


***Supp Fig.4:*** **Dorso Lateral Prefrontal Cortex (DLPFC) transcriptome trajectories across development**. On the x-axis the developmental stages with the vertical dashed line representing the birth. PCW = Post Conception Weeks, mos = months and y = years. On the y-axis the normalized expression averaged across the Bonferroni significant genes.

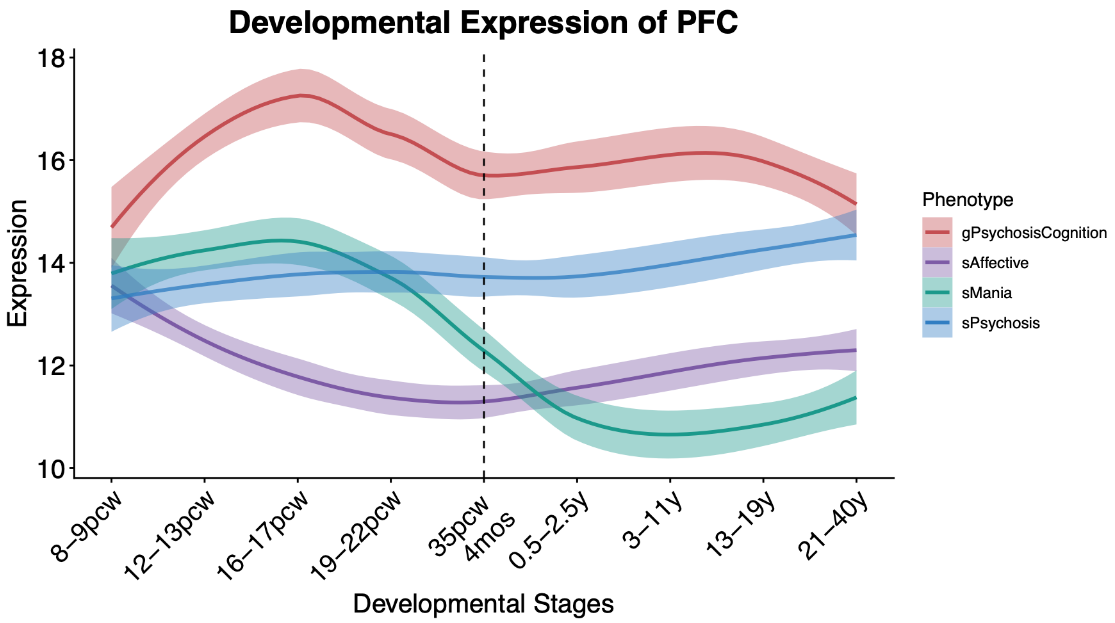


***Supp Fig.5:*** **Prefrontal Cortex (PFC) transcriptome trajectories across development**. On the x-axis the developmental stages with the vertical dashed line representing the birth. PCW = Post Conception Weeks, mos = months and y = years. On the y-axis the normalized expression averaged across the Bonferroni significant genes.

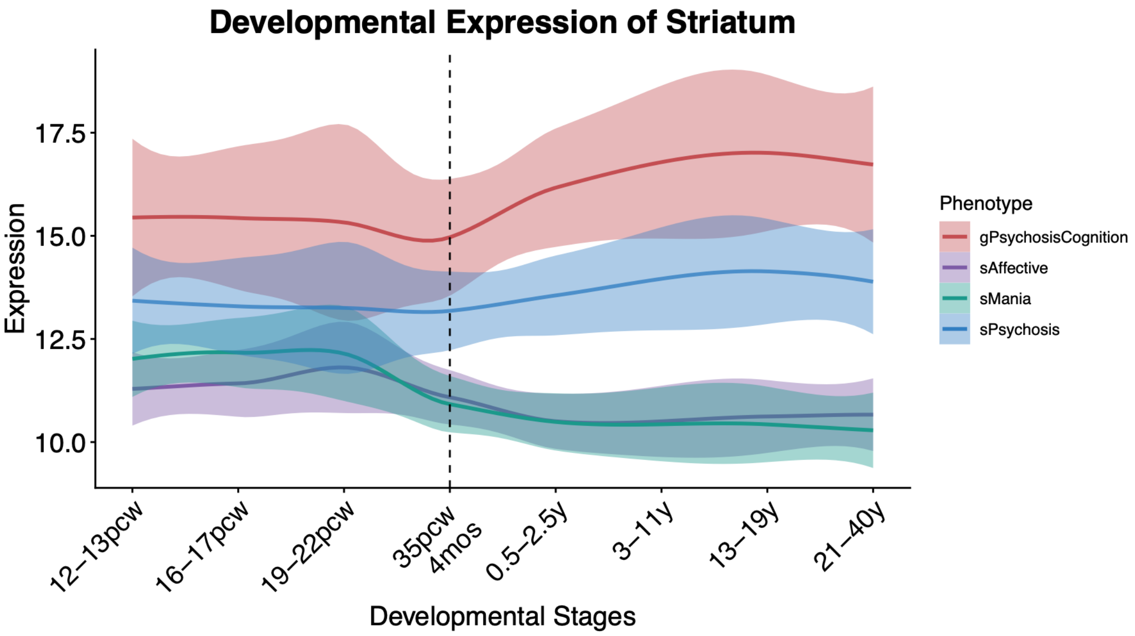


***Supp Fig.6:*** **Striatum transcriptome trajectories across development**. On the x-axis the developmental stages with the vertical dashed line representing the birth. PCW = Post Conception Weeks, mos = months and y = years. On the y-axis the normalized expression averaged across the Bonferroni significant genes.
